# Using human genetics to understand the causes and consequences of circulating cardiac troponin I in the general population

**DOI:** 10.1101/2020.09.04.20187401

**Authors:** Marta R Moksnes, Helge Røsjø, Anne Richmond, Magnus N Lyngbakken, Sarah E Graham, Ailin Falkmo Hansen, Brooke N Wolford, Sarah A Gagliano Taliun, Jonathon LeFaive, Humaira Rasheed, Laurent F Thomas, Wei Zhou, Nay Aung, Ida Surakka, Nicholas J Douville, Archie Campbell, David J Porteous, Steffen E. Petersen, Patricia B. Munroe, Paul Welsh, Naveed Sattar, George Davey Smith, Lars G Fritsche, Jonas B Nielsen, Bjørn Olav Åsvold, Kristian Hveem, Caroline Hayward, Cristen J Willer, Ben M Brumpton, Torbjørn Omland

**Affiliations:** K.G. Jebsen Center for Genetic Epidemiology, Department of Public Health and Nursing, NTNU - Norwegian University of Science and Technology, Trondheim, Norway; Division of Research and Innovation, Akershus University Hospital, Lørenskog, Norway; Institute of Clinical Medicine, Faculty of Medicine, University of Oslo, Oslo, Norway; MRC Human Genetics Unit, Institute of Genetics and Molecular Medicine, University of Edinburgh, Edinburgh, UK; Department of Cardiology, Division of Medicine, Akershus University Hospital, Lørenskog, Norway; Division of Cardiovascular Medicine, Department of Internal Medicine, University of Michigan, Ann Arbor, MI, USA; Department of Computational Medicine and Bioinformatics, University of Michigan, Ann Arbor, MI, USA; Faculty of Medicine, Université de Montréal, Montréal, Québec, Canada; Montréal Heart Institute, Montréal, Québec, Canada; Department of Biostatistics, University of Michigan School of Public Health, Ann Arbor, MI, USA; Center for Statistical Genetics, University of Michigan School of Public Health, Ann Arbor, MI, USA; MRC Integrative Epidemiology Unit, University of Bristol, Bristol, UK; Department of Clinical and Molecular Medicine, NTNU - Norwegian University of Science and Technology, Trondheim, Norway; BioCore - Bioinformatics Core Facility, NTNU - Norwegian University of Science and Technology, Trondheim. Norway; Clinic of Laboratory Medicine, St. Olavs hospital, Trondheim University Hospital, Trondheim, Norway; Analytic and Translational Genetics Unit, Massachusetts General Hospital, Boston, Massachusetts, USA; Program in Medical and Population Genetics, Broad Institute of Harvard and MIT, Cambridge, Massachusetts, USA; Stanley Center for Psychiatric Research, Broad Institute of Harvard and MIT, Cambridge, Massachusetts, USA; William Harvey Research Institute, Queen Mary University of London, Charterhouse Square, London EC1M 6BQ, UK; National Institute for Health Research Barts Cardiovascular Biomedical Research Centre, Queen Mary University of London, London, UK; Barts Heart Centre, St. Bartholomew’s Hospital, Barts Health NHS Trust, West Smithfield, London EC1A 7BE, UK; Department of Anesthesiology, University of Michigan, Ann Arbor, MI, USA 19; Medical Genetics Section, CGEM, Institute of Genetics and Molecular Medicine, University of Edinburgh, Edinburgh, UK; Institute of Cardiovascular and Medical Sciences, University of Glasgow, Glasgow, UK; Department of Epidemiology Research, Statens Serum Institute, Copenhagen, Denmark; Department of Cardiology, Copenhagen University Hospital, Copenhagen, Denmark; HUNT Research Centre, Department of Public Health and Nursing, NTNU - Norwegian University of Science and Technology, Levanger, Norway; Department of Endocrinology, St. Olavs hospital, Trondheim University Hospital, Trondheim, Norway; Department of Human Genetics, University of Michigan, Ann Arbor, MI, USA; Clinic of Thoracic and Occupational Medicine, St. Olavs hospital, Trondheim University Hospital, Trondheim, Norway

**Author notes:** These authors contributed equally to this work (Marta R Moksnes, Helge Røsjø, Anne Richmond). These authors contributed equally to this work (Caroline Hayward, Cristen J Willer, Ben M Brumpton, Torbjørn Omland).

## Abstract

Circulating cardiac troponin proteins are associated with structural heart disease and predict incident cardiovascular disease in the general population. However, the genetic contribution to cardiac troponin I (cTnI) concentrations and its causal effect on cardiovascular phenotypes is unclear. We combine data from two large population-based studies, the Trøndelag Health Study and the Generation Scotland Scottish Family Health Study and perform a genome-wide association study of high-sensitivity cTnI concentrations with 48 115 individuals. We further used two-sample Mendelian randomization to investigate the causal effects of circulating cTnI on acute myocardial infarction and heart failure.

We identified 12 genetic loci (8 novel) associated with cTnI concentrations. Associated protein-altering variants highlighted putative functional genes: *CAND2, HABP2, ANO5, APOH, FHOD3, TNFAIP2, KLKB1* and *LMAN1*. Phenome-wide association tests in 1283 phecodes and 274 continuous traits in UK Biobank showed associations between a polygenic risk score for cTnI and cardiac arrhythmias, aspartate aminotransferase 1 and anthropometric measures. Excluding individuals with a known history of comorbidities did not materially change associations with cTnI. Using two-sample Mendelian randomization we confirmed the non-causal role of cTnI in acute myocardial infarction (5 948 cases, 355 246 controls). We found some indications for a causal role of cTnI in heart failure (47 309 cases and 930 014 controls), but this was not supported by secondary analyses using left ventricular mass as outcome (18 257 individuals).

Our findings clarify the biology underlying the heritable contribution to circulating cTnI and support cTnI as a non-causal biomarker for acute myocardial infarction and heart failure development in the general population. Using genetically informed methods for causal inference of cTnI helps inform the role and value of measuring cTnI in the general population.

## Introduction

Cardiac specific troponins I (cTnI) and T (cTnT) are structural proteins involved in cardiac muscle contraction. During acute myocardial ischemia, cardiac troponin proteins are released into the blood stream and are used to diagnose acute myocardial infarction (AMI)(1). In AMI, cardiac troponin release is a result of direct ischemic cardiomyocyte injury. In contrast, the pathobiology of cardiac troponin release outside of AMI is not known, although low-level cardiac troponin elevations are associated with future cardiovascular events in the general population, leading some clinicians to suggest a need for wider use of cardiac troponins in risk stratification in general(2). Hence, a better understanding of low-level cardiac troponin release is needed, and the study of large population-based cohorts with genetic data could bring new insights.

The genetic determinants of cardiac troponin in the general population have not been studied in detail, and the downstream biology, including possible causal effects on cardiovascular disease, is unclear. The cTnI and cTnT proteins are encoded by the *TNNI3* and *TNNT2* genes(3), respectively, and mutations in these genes are recognized as disease-causing mutations in patients with familial cardiomyopathies(4,5). Patients with cardiomyopathies demonstrate increased concentrations of circulating cardiac troponins(6), and these conditions are strongly associated with heart failure (HF) development(7). We recently demonstrated a close association between cTnI and left ventricular hypertrophy(8), which represents an intermediate stage prior to symptomatic HF(9). Moreover, as experimental studies have found immune responses targeting cTnI to aggravate acute cardiac damage and to induce myocardial dysfunctions and HF(10,11), cTnI may have a direct or indirect causal effect on HF development in humans. Circulating cardiac troponin concentrations are also influenced by protein degradation and clearance(12). A previous genome-wide association study (GWAS) in the general population from the Generation Scotland Scottish Family Health Study (GS:SFHS) reported five genetic loci associated with cTnI concentrations and four other loci associated with cTnT concentrations(13), however without replication in an independent cohort. More knowledge about the genetic determinants of circulating cardiac troponins could reveal new pathobiology important for HF development. In contrast, as AMI type 1 is a result of an acute epicardial coronary artery occlusion and cTnI is only expressed in cardiac myocytes, no causal role for cTnI is expected for incident AMI. Due to limited knowledge of the genetic contribution to circulating cardiac troponins, no previous studies have used genetically informed methods of causal inference, namely Mendelian randomization (MR), to assess the causal effect of cardiac troponin concentrations on AMI or HF(14).

High-sensitivity cTnI has been cross-sectionally and repeatedly measured in three surveys of the Trøndelag Health Study(15) (HUNT) from 1995 to 2019. Additionally, recent efforts to sequence 53 831 diverse genomes in the NHLBI TOPMed Program(16) have enabled the imputation of more than 26 million variants in HUNT and GS:SFHS combined, including less common loss-of-function variants that have not been previously investigated(13).

In order to validate previous findings and discover novel genetic variants associated with circulating cTnI in the general population, we conducted a genome-wide meta-analysis of cTnI concentration in 48 115 individuals from HUNT and GS:SFHS. Additionally, we assessed changes in cTnI with time in 5 178 HUNT participants with repeated measures. Finally, we performed MR to investigate the causal effects of circulating cTnI on AMI and HF.

## Methods

### Cohort Descriptions

The recruitment and design of the HUNT and GS:SFHS studies have been reported elsewhere(15,17). Participants (aged ≥ 20 years in HUNT and 35–65 years in GS:SFHS) have been genotyped using Illumina HumanCoreExome array 12 v.1.0 or v.1.1 in HUNT and Illumina HumanOmniExpressExome-8 v1_A or v1-2 in GS:SFHS. Sample and variant quality control (QC) was performed using standard practices and has been reported in detail for both studies elsewhere(18,19). All variants were imputed from the TOPMed reference panel(16) (freeze 5) using Minimac4 v1.0 (see Web Resources). The reference panel is composed of 53 831 multi-ethnic samples and 308 107 085 single nucleotide variants (SNVs) and indel variants at high depth (mean read depth 38.2X). The individuals included in the analyses were of European ancestry.

### Cardiac Troponin I

High-sensitivity cTnI concentrations (Architect STAT, Abbott Laboratories, Abbott Diagnostics) were measured from serum samples in both studies using the manufacturers’ protocol and quality assurance scheme.

In HUNT, non-fasting serum samples were drawn in 1995-1997 (HUNT2), 2006-2008 (HUNT3) and 2017-2019 (HUNT4). The HUNT2 and HUNT3 samples were centrifuged at room temperature and stored at -80°C after collection. The HUNT2 samples were thawed and refrozen once in 2008 and subsequently stored at -20°C. High-sensitivity cTnI concentrations were analyzed in 2014 from HUNT2, in 2015 from HUNT3 and was analyzed consecutively in 2017-2019 from fresh serum samples in HUNT4. We combined cTnI measurements from all studies into one variable for cTnI concentration: If an individual had measurements from multiple HUNT surveys, we used the earliest measurement in the analyses, assuming participants to be less exposed to potential confounding factors at a younger age. After prioritization, 29 839 individuals of European ancestry had measurements of cTnI in HUNT, of which 23 990 were above or equal to the lower limit of detection (1.2 ng/L) reported for the assay(20). Measurements below 1.2 ng/L are imprecise on an individual level but were included since they collectively could add information to the GWAS analysis.

We defined change in cTnI concentration per time as the difference in cTnI concentration from HUNT2 to HUNT3 divided by the follow-up time. The median follow-up time was 10 years and 7 months. In total, 5 178 participants had data of change in cTnI over time.

In GS:SFHS, fasting blood samples were collected between 2006 and 2010. Serum samples were centrifuged, and serum aliquots stored at –80°C until future biochemical analyses. A total of 18 276 individuals had measurements of cTnI in GS:SFHS.

Measurements below the reported limit of detection (1.2 ng/L)(20) were reported as half of the limit of detection (0.6 ng/L) since the original measurements were unavailable.

### Association Analyses

We used a linear mixed model regression under an additive genetic model for each variant as implemented in BOLT-LMM (v.2.3.4)(21) to control for relatedness between the samples in all analyses.

We included age, sex, genotyping batch and the first 10 genetic principal components of ancestry (PCs) as covariates in both HUNT and GS:SFHS. Additionally, we included as a covariate in HUNT the survey from which the serum sample was collected for cTnI measurement. We applied rank-based inverse normal transformation for the cTnI concentration after adjusting for age and sex using linear regression (additionally adjusting for survey in HUNT). Variants with a minor allele count (MAC) < 10 or imputation R^2^ < 0.3 were excluded, leaving 21 233 433 variants in the HUNT analysis and 21 677 569 variants in the GS:SFHS analysis. We performed genomic control correction after each analysis.

### Meta-analysis

We performed a fixed-effect inverse-variance weighted meta-analysis of 26 711 992 variants imputed in up to 48 115 individuals using METAL(22). We considered genetic loci reaching a p-value < 5×10^−8^ in the meta-analysis for follow-up analysis.

### Sensitivity Analyses

To investigate if the identified genetic loci could be explained by clinical endpoints associated with chronically elevated cardiac troponin levels, we performed a sensitivity analysis of cTnI concentration in HUNT, excluding 5 253 individuals with a current or known history of comorbidities (atrial fibrillation [ICD9 427.3, ICD10 I48], diabetes mellitus [self-reported], HF[ICD9 428, ICD10 I50], cardiomyopathy [ICD9 425, ICD10 I42] or impaired kidney function [estimated glomerular Filtration rate [eGFR] < 60 ml/min/1.73m^2^] in the HUNT survey where the serum sample was collected for cTnI measurement).

### Heritability Estimation

We estimated the narrow-sense (additive) SNV heritability of cTnI concentration in HUNT using genome-wide complex trait analysis (GCTA) (23,24). We created a genetic relationship matrix (GRM) based on all genotyped autosomal variants in the 29 839 HUNT participants with troponin measurements and used the GRM with GCTA-GREML (genomic-relatedness-based restricted maximum-likelihood) to estimate the variance in cTnI concentration explained by the variants. Age, sex, genotyping batch and HUNT survey were used as covariates in the analysis, and the cardiac troponin measurement was transformed to normality with rank based inverse normal transformation after regression on age, sex and HUNT survey prior to analysis.

### Variant Annotation and Fine-mapping

We annotated genetic variants using ANNOVAR (v.2019Oct24)(25). For fine-mapping of the cTnI loci we applied SuSiE (v0.7.1)(26), which uses iterative Bayesian stepwise selection to estimate the number of causal variants in each locus and create credible sets with a 95% cumulative posterior probability for containing a causal variant. The LD matrix used with SuSiE was calculated from the imputed variants of 5 000 unrelated individuals in HUNT, and variants only imputed in GS:SFHS were therefore excluded from the fine-mapping. We visualized the fine-mapped regions with LocusZoom(27).

### Tissue, Gene and Gene Set Enrichment Tests

We performed gene set enrichment, gene prioritization and tissue/cell type enrichment tests on the cTnI loci using DEPICT (v1.0)(28). To ensure that DEPICT could identify variants in all loci we filtered the meta-analysis results to the 3 906 791 variants overlapping with the DEPICT reference files. We used plink (v1.9)(29) to identify two sets of independent variants by clumping the filtered results into 500 kb regions of correlated variants (r^2^ < 0.05) meeting two different p-value cut-offs (p-value < 1×10^−5^ and p-value < 5×10^−8^). The two sets of independent variants were used as input for DEPICT and analyzed separately. All cTnI loci were included in the analyses except variants in the HLA region.

### Expression Quantitative Trait Locus Tests

To identify cTnI index variants associated with the expression level of a gene in specific tissue types, we downloaded single tissue cis-eQTL data from the Genotype-Tissue Expression (GTEx) portal, data set v8 (see Web Resources). We identified all significant associations (p-value < 1×10^−9^ [5×10^−8^/49 tissues]) between cTnI index variants and gene expression levels in any tissue type. To further assess if any cis-eQTL signals and GWAS meta-analysis signals were consistent with a shared causal variant for cTnI concentration and gene expression, we used Bayesian colocalization analysis as implemented in the R package coloc(30). For each tissue type, we analyzed all genes whose expression was associated (p-value < 5×10^−8^) with at least one cTnI associated variant (p-value < 5×10^−8^), using effect sizes and standard errors for each variant-trait association as input. The coloc software estimated the variance in each trait (cTnI concentration or gene expression level) from the sample sizes and minor allele frequencies. We set the prior probability of a genetic variant being associated with only cTnI concentrations, only gene expression or both traits to be 10^−4^, 10^−4^ and 10^−6^ respectively. We considered posterior probabilities (PP4) above 75% to give strong support for a common causal variant for the two traits in a given tissue, posterior probability between 50% and 75% to be inconclusive or to give weak support for a common causal variant, and posterior probabilities < 50% to indicate no support for a common causal variant between the two traits.

### Polygenic Risk Score for cTnI

We used R (v3.6.2)(31) to construct a polygenic risk score (PRS) for cTnI, by summing the product of the effect size and the estimated allele count for the 12 index variants in genome-wide significant loci (p-values < 5×10^−8^). To test the association between the PRS and cTnI in an independent sample, we performed the GWAS on cTnI concentrations in 26 415 HUNT participants, excluding 3 424 individuals who were unrelated (no 3rd degree relatives or closer) both to each other and to the individuals in the GWAS. We used KING (v2.1.5)(32) to estimate the relatedness. The smaller GWAS in HUNT (N = 26 415) was meta-analyzed with the full GWAS from GS:SFHS to obtain effect estimates independent of the excluded individuals. We constructed the PRS from the effect size estimates from this meta-analysis and used a linear regression model to test the association between the PRS and serum concentrations of cTnI (inverse rank transformed to normality) in the independent sample of 3 424 unrelated HUNT participants.

### Phenome-wide Association Tests (PheWAS)

We tested the association of the PRS (PRS-PheWAS), as well as the individual index variants (single variant PheWAS), with 1 688 phecodes and 83 continuous biomarkers and traits in participants of white British ancestry in UK Biobank. We calculated a PRS for each participant as described in the above paragraph, using TOPMed imputed estimated allele counts and effect sizes from the meta-analysis. We used a logistic or linear regression model respectively to assess the association of the single variant estimated allele counts or inverse normalized PRS and each phecode or continuous biomarker. For the PRS-pheWAS we included as covariates sex and birth year for binary traits, and sex and age at measurement for continuous traits. For the single variant pheWAS we used GWAS summary statistics generated with SAIGE (v.29.4.2)(33), with sex and the first four PCs as covariates in addition to age at initial assessment for quantitative traits and birth year for binary traits. To correct for multiple testing of phecodes and continuous variables we used a Bonferroni corrected significance cut-off of 2.8×10^−5^.

### Mendelian Randomization of cTnI on AMI and HF

To assess causal associations of circulating cTnI on AMI or HF, we performed inverse variance weighted (IVW) two-sample MR analyses using R (v3.6.2)(31) with the TwoSampleMR(34) and MRInstruments(35) libraries. We used the effect sizes of 11 index variants from the meta-analysis as genetic instrument for cTnI, while we found the respective associations and effect sizes of the instrument variants for AMI (ICD10 I21) and HF (ICD10 I50) from independent genome-wide summary level data (5 948 AMI cases vs 355 246 controls from UK Biobank(36) available from the Neale laboratory (see Web Resources) and 47 309 HF cases vs 930 014 controls from the Heart Failure Molecular Epidemiology for Therapeutic Targets consortium [HERMES] consortium(37) available from the Cardiovascular Disease Knowledge Portal (see Web Resources). The rare cTnI index variant rs151313792 was excluded from both analyses because it was not present in either outcome data set, and we were unable to identify a proxy variant. In the analysis of HF, the variant rs8039472 was used as a proxy for rs8024538 (r^2^_HUNT_ = 0.98) because rs8024538 was not present in the HF data. We performed sensitivity analyses using Steiger filtered IVW, weighted mode, weighted median, penalized weighted median and MR Egger. To evaluate the reverse causal association of HF on cTnI concentration, we also performed an IVW two-sample MR analysis using the effect sizes of 11 index variants for HF, and the respective associations and effect sizes of the same variants from our GWAS meta-analysis of cTnI. We further performed sensitivity analyses using left ventricular mass, which is associated with HF development. We found the associations and effect sizes of these variants for left ventricular mass from an independent GWAS in 18 257 participants in UK Biobank(38).

### Ethics

Participation in HUNT is based on informed consent, and the study has been approved by the Norwegian Data Protection Authority and the Regional Committee for Medical and Health Research Ethics in Central Norway (REK Reference number: 2015/2294).

The GS:SFHS obtained written informed consent from all participants and received ethical approval from the National Health Service Tayside Committee on Medical Research Ethics (REC Reference number: 05/S1401/89).

This study was covered by the ethics approval for UK Biobank studies (application 24460) from the NHS National Research Ethics Service on 17th June 2011 (Ref 11/NW/0382) and extended on 10th May 2016 (Ref 16/NW/0274).

## Results

### Discovery of Genetic Loci Associated with cTnI

After a discovery genome-wide association meta-analysis of HUNT (n = 29 839) and GS:SFHS (n = 18 276), we identified 12 genetic loci associated (p-value < 5×10^−8^) with cTnI concentrations (Tables 1, S1, S2, Figures S1, S2). All 12 loci were well imputed in both HUNT and GS:SFHS and had consistent directions and effect sizes across both studies. Eight of the 12 loci were previously unreported, while four were previously reported, but not yet replicated(13). Additionally, we discovered 11 novel, rare variants (MAF < 0.001, p-value < 5×10^−8^) that were only imputed in HUNT and could not therefore be replicated (Table S1). Excluding individuals with a known history of comorbidities did not materially change the results (Table S4). No genetic loci were genome-wide significant for changes in cTnI concentrations with time, however three loci were suggestive (p-value < 10^−6^) (Tables S3, S5).

**Table 1.** Index variants in genome-wide significant cTnI loci

| Novelty | rsID | Chr:Pos:Ref:Alt | Consequence | Reference Gene | AF | Effect Size | SE | P-value |
| --- | --- | --- | --- | --- | --- | --- | --- | --- |
| Novel | rs7650482 | 3:12800305:A:G | intronic, $r^2 = 0.91$ with protein-altering variant | CAND2 | 0.64 | 0.04 | 0.01 | $4 \times 10^{-10}$ |
| Known | rs12331618 | 4:186218785:A:G | intergenic; $r^2 = 1.00$ with protein-altering variant | CYPV4;KLKB1 | 0.52 | 0.04 | 0.01 | $2 \times 10^{-12}$ |
| Novel | rs2731672 | 5:177415473:T:C | intergenic | F12;GRK6 | 0.74 | 0.05 | 0.01 | $9 \times 10^{-14}$ |
| Novel | rs9391746 | 6:32715903:G:T | intergenic | HLA-DQB1;<br>HLA-DQA2 | 0.07 | 0.08 | 0.01 | $1 \times 10^{-8}$ |
| Known | rs2270552 | 10:74103992:C:T | intronic | VCL | 0.68 | -0.06 | 0.01 | $3 \times 10^{-19}$ |
| Novel | rs7080536 | 10:113588287:G:A | protein-altering | HABP2 | 0.04 | -0.10 | 0.01 | $1 \times 10^{-12}$ |
| Known | rs7481951 | 11:22250324:A:T | protein-altering | ANO5 | 0.59 | 0.05 | 0.01 | $2 \times 10^{-19}$ |
| Novel | rs4283165 | 14:103137158:G:A | 3' UTR, $r^2 = 0.65$ with protein-altering variant | TNFAIP2 | 0.65 | 0.04 | 0.01 | $7 \times 10^{-9}$ |
| Novel | rs8024538 | 15:84825188:A:T | intronic | ALPK3 | 0.53 | 0.04 | 0.01 | $7 \times 10^{-9}$ |
| Novel | rs1801690 | 17:66212167:C:G | protein-altering | APOH | 0.06 | 0.10 | 0.01 | $1 \times 10^{-14}$ |
| Novel | rs151313792 | 18:36681510:G:A | protein-altering | FHOD3 | 0.004 | 0.26 | 0.05 | $1 \times 10^{-8}$ |
| Known | rs12604424 | 18:59353974:T:C | Intronic, $r^2 = 1.00$ with protein-altering variant | LMAN1 | 0.13 | 0.06 | 0.01 | $1 \times 10^{-11}$ |

The variants are given with chromosome (Chr), genomic position in GRCh38 (Pos), reference (Ref) and alternate (Alt) alleles. Consequence, frequency (AF) and effect size with standard error (SE) are given for the alternate allele.

We found three protein-altering index variants in cTnI loci: rs7080536 (MAF = 0.04, effect size = -0.10, p-value = 1×10^−12^) in *HABP2*, rs1801690 (MAF = 0.06, effect size = 0.10, p-value = 1×10^−14^) in *APOH*, and rs7481951 (MAF = 0.41, effect size = 0.05, p-value = 2×10^−19^) in *ANO5*. Furthermore, we identified a rare protein-altering index variant (rs151313792, MAF = 0.004, effect size = 0.26, p-value = 1×10^−8^) in *FHOD3*. Two additional index variants were in perfect linkage disequilibrium (LD) (r^2^=1.00) with protein-altering variants in *KLKB1* and *LMAN1*, as described in the section on fine-mapping below.

### Fine-mapping for Causal Variants in the cTnI Loci

We identified 14 credible sets (Table S9) of genetic variants in the cTnI loci using SuSiE. For each credible set there was a 95% cumulative posterior probability that it contained at least one causal variant. The sizes of the credible sets ranged from 1 variant to 64 variants (in the HLA region and one region on chromosome 10), providing varying evidence for the importance of specific variants. Nine of the cTnI index variants (rs7650482, rs12331618, rs7080536, rs7481951, rs4283165, rs8024538, rs1801690, rs151313792 and rs12604424) were found in a credible set, and one of these credible sets only contained the index variant itself (rs7080536), suggesting that this was the most likely causal variant in the locus. The two loci with the lowest p-values (chromosomes 10 and 11) gave rise to two credible sets each, indicating multiple independent signals. However, the credible sets were in close proximity or overlapping and therefore the seemingly independent signals might have resulted from uncaptured LD structures.

Annotation of the variants in each credible set (Table S10) revealed four additional protein-altering variants in moderate to high LD with non-exonic index variants (Table 1) : rs11718898 in *CAND2*, correlation r^2^ in HUNT (r^2^_HUNT_) = 0.91 with index variant rs7650482 (Figure 1a); rs3733402 in *KLKB1*, r^2^_HUNT_ = 1.00 with index variant rs12331618 (Figure 1b); rs1132339 in *TNFAIP2*, r^2^_HUNT_ = 0.65 with index variant rs4283165 (Figure 1c); rs2298711 in *LMAN1*, r^2^_HUNT_ = 1.00 with index variant rs12604424 (Figure 1d). The rs2298711 protein-altering variant was reported as index variant in the previous GWAS study in GS:SFHS(13).

**Figure 1:**
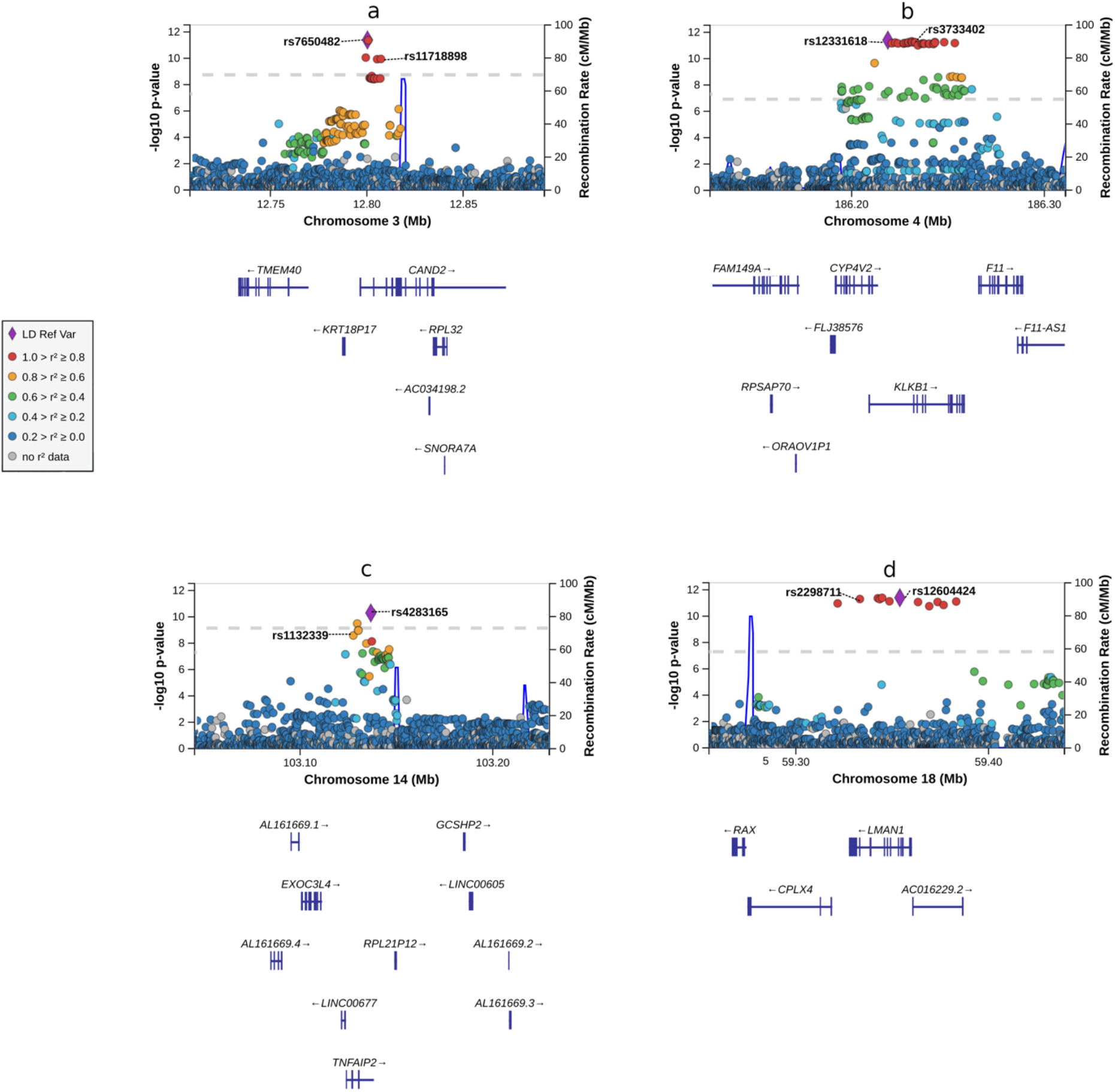
LocusZoom plots of cTnI loci with protein-altering variants in moderate or high LD with the index variant: Chromosome positions (GRCh38) are given on the x-axes, and the meta-analysis -log10(p-value) for the variants are given on the y-axis. T The genome-wide significance p-value threshold (5 x 10^−8^) is indicated by the dotted line. The correlation r^2^ for each variant is indicated by colors relative to the index variants (LD Ref Var) in each panel: Panel a: The CAND2 locus on chromosome 3 with index variant rs7650482. A protein-altering variant in CAND2, rs11718898 (pVal77Ala), is in high LD with the index variant. Panel b: The KLKB1 locus on chromosome 4 with index variant rs12331618. A protein-altering variant in KLKB1, rs3733402 (p.Ser143Asn, p.Ser105Asn), is in perfect LD with the index variant. Panel c: The TNFAIP2 locus on chromosome 14 with index variant rs4283165. A protein-altering variant in TNFAIP2, rs1132339 (p.Gln282Glu), is in moderate LD with the index variant. Panel d: The LMAN1 locus on chromosome 18 with index variant rs12604424. A protein-altering variant in LMAN1, rs2298711 (p.Met410Leu), is in perfect LD with the index variant.

Furthermore, we found a rare exonic frameshift substitution (rs137854521 in *ANO5*, r^2^_HUNT_ = 0.003, D’ = 1.00 with the more common index variant rs7481951) in the second credible set in the same region as rs7481951, and a variant in another credible set was located in the *F12* 5’ untranslated region (UTR).

### Prioritization of Genes, Pathways and Tissues

To further explore the underlying biology of the cTnI associated loci, we used DEPICT to identify candidate causal genes at each locus. For suggestive (p-value < 1×10^−5^) cTnI loci (excluding the HLA region), we identified gene sets enriched for genes in cTnI loci, prioritized genes in each locus that shared predicted function with the genes in the remaining cTnI loci more often than expected by chance, and identified tissues and cell types in which cTnI locus genes were highly expressed (Tables S6-S8). After removing results with false discovery rate above 5%, we found an enriched expression of suggestive cTnI genes in heart and heart ventricle tissues, and in tooth development. Among 14 461 reconstituted gene sets, one gene set (“abnormal placenta morphology”) was enriched with genes in significant cTnI loci. DEPICT prioritized the genes *KLHL21, THAP3, CAND2, AOPEP, CTNNA3, SYNPO2L, VCL, ANXA11, MYOF, NRAP, CASP7, ANO5, ALPK3* and *ZNF592* based on shared predicted functions with genes from the other associated loci. Most of these (*CAND2, VCL, ANO5, ALPK3, NRAP, CASP7, SYNPO2L, ZNF592*) were in significant (p-value < 1×10^−8^) cTnI loci, but not always near the index variant.

### Heritability of cTnI Concentration

We estimated the proportion of phenotypic variance explained by all genotyped autosomal variants using GCTA and found a narrow-sense SNV heritability of cTnI concentration in HUNT (variance explained, V_g_/V_p_ ± 1SE) to be 0.15 ± 0.01, p-value 2.63×10^−32^. The HUNT cohort has a high degree of relatedness and is therefore more genetically homogeneous than cohorts with fewer relatives. Including more rare variants from the TOPMed panel would further increase the homogeneity of the dataset. This might, in part, explain the lower heritability observed in HUNT than in previous studies(13,39).

### Variants Associated with Tissue-Specific cis-eQTLs

Five of the cTnI index variants were significantly associated with the expression of one or more genes in at least one tissue type (Table S11). Two of these variant-gene associations were found in the cardiovascular system: rs7481951 was associated with the expression of *ANO5* in aortic and tibial arteries and rs12331618 was associated with the expression of *ADK* in tibial arteries.

To discover shared causal variants for cTnI concentration and expression levels of nearby genes in different tissues, we used the cis-eQTL data from GTEx v8 and performed a Bayesian colocalization analysis of the gene expression signals and cTnI concentration signals. Four cTnI loci were significantly colocalized (posterior probability of common causal variant > 75%) with the cis-eQTL signals for a gene in at least one tissue (Table S12): The expression of coagulation factor *F11* in tibial arteries (rs12331618), the expression of coagulation factor *F12* in liver (rs2731672), the expression of *BMS1P4* (pseudogene) in whole blood (rs2270552), the expression of *ADK* in the sigmoid colon (rs2270552), as well as the expression of lncRNA gene *Lnc*-*WDR73*-10 in the left heart ventricle, mammary breast tissue and brain frontal cortex (rs8024538). The association signal for *ADK* was also near the posterior probability cut-off of 75% in aortic tissue (rs2270552) (Table S12).

### PheWAS of cTnI Loci and cTnI Polygenic Risk Score

Four of the index variants associated with increased cTnI concentrations were significantly associated with diseases or continuous traits in UK Biobank (Figures 2 and 3, Tables S13, S14) after correction for multiple testing: The variant rs7650482 was associated with cardiac arrhythmias, in particular atrial fibrillation and flutter, as well as waist hip ratio, standing height and basal metabolic rate. In addition, the variant rs12331618 was associated with pulmonary heart disease and phlebitis/thrombophlebitis, the variant rs2731672 was associated with standing height and basal metabolic rate, and the variant rs9391746 was mainly associated with autoimmune inflammatory conditions.

**Figure 2:**
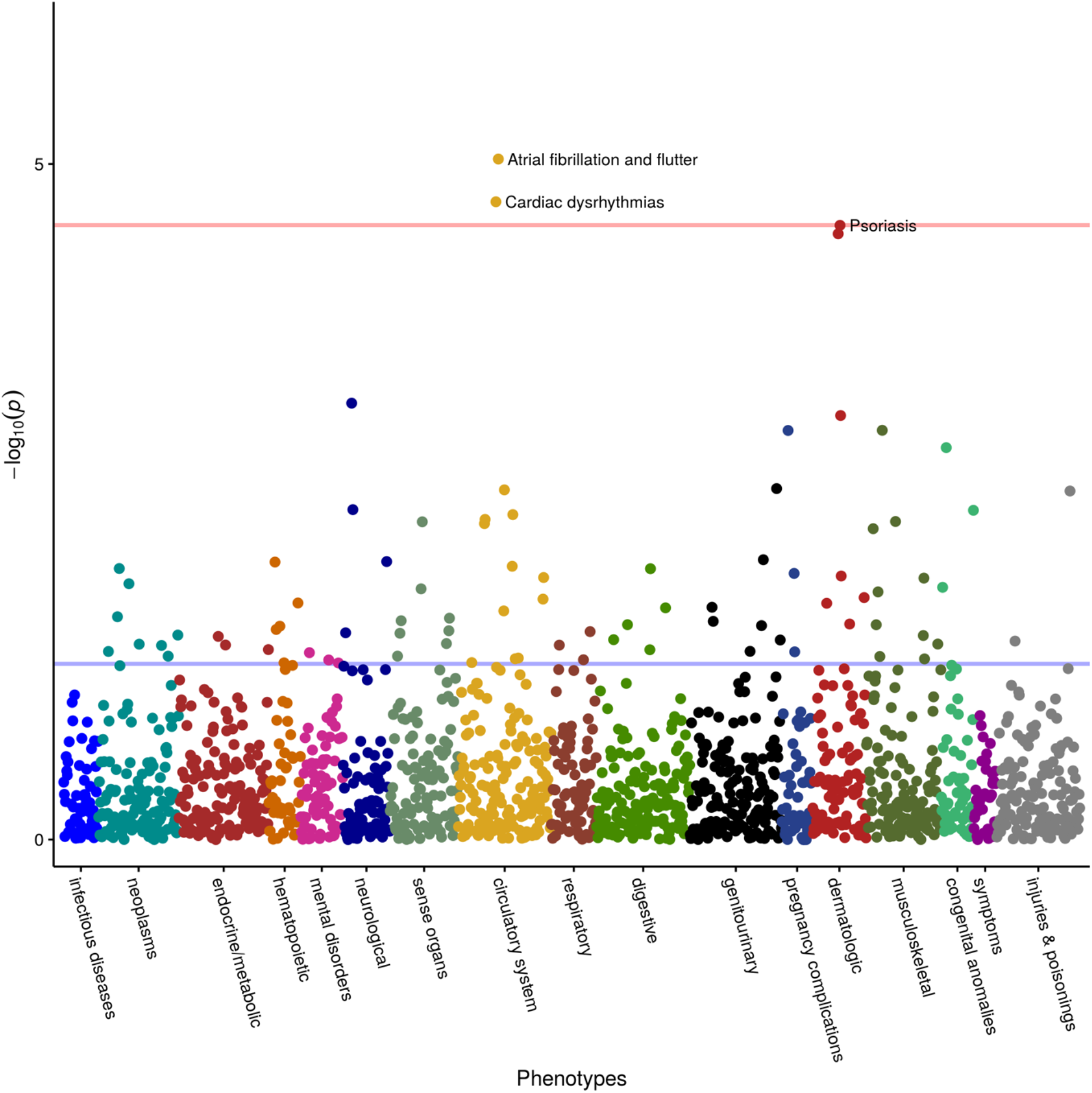
PRS-PheWAS: Associations of PRS (increasing cTnI concentrations) with 1 688 phecodes in UK Biobank.

**Figure 3:**
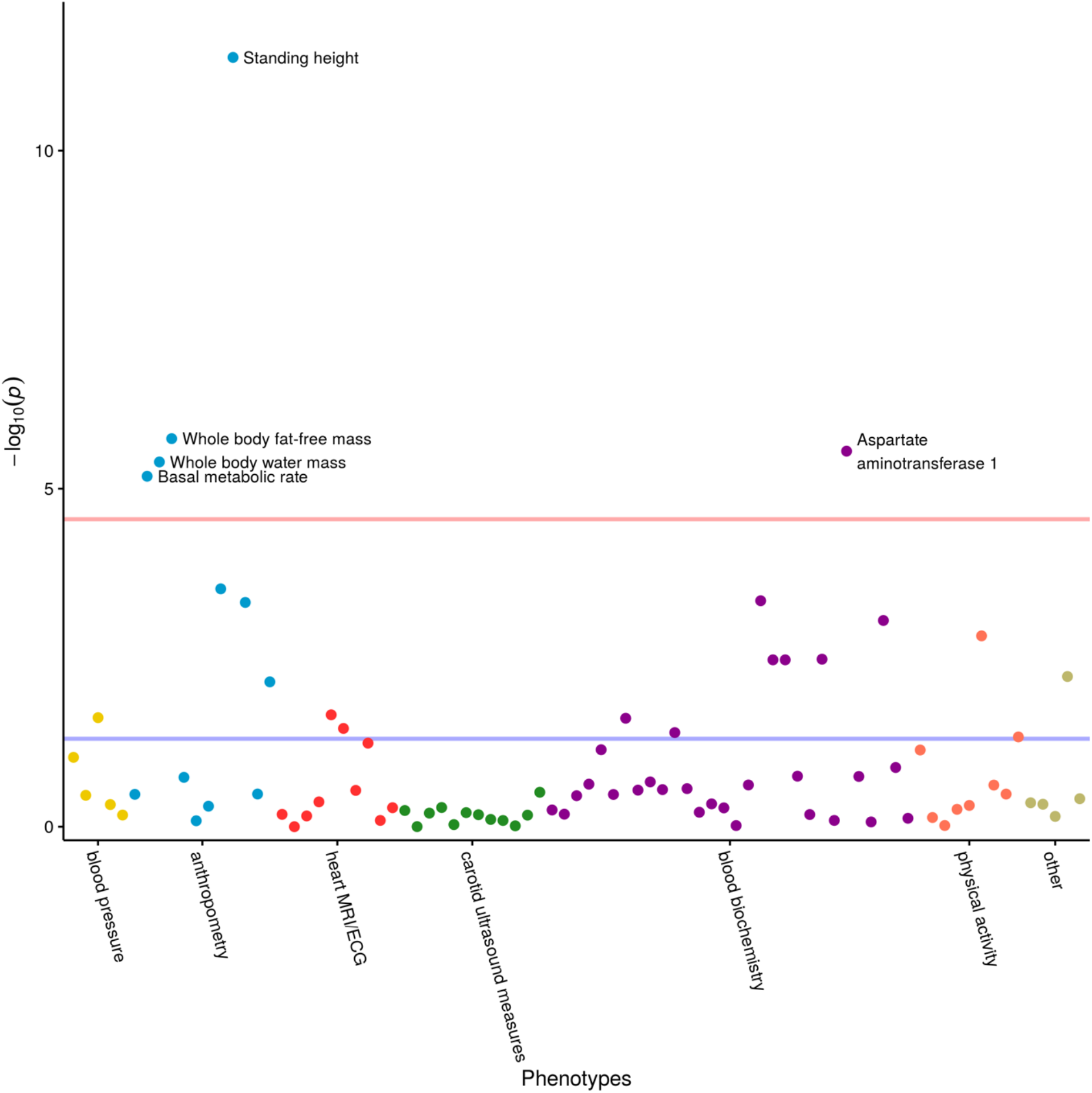
PRS-PheWAS: Associations of PRS (increasing cTnI concentrations) with 83 continuous traits and biomarkers in UK Biobank. ECG: Electrocardiogram, MRI: Magnetic resonance imaging.

The polygenic risk score (PRS) based on genome-wide significant index variants (n = 12, variance explained=1.2%) was associated with seven phecodes and continuous traits in UK Biobank after correction for multiple testing (Figures 2, 3, Tables S15, S16): cardiac arrhythmias in general, and the sub-phenotype atrial fibrillation and flutter in particular, as well as standing height, aspartate aminotransferase 1, whole body fat free mass, whole body water mass and basal metabolic rate.

### Mendelian Randomization

To investigate the causal effect of circulating cTnI on both AMI and HF, we performed two-sample MR using 11 of 12 index variants from our meta-analysis as an instrument for cTnI (F-statistic = 44, r^2^ = 0.012 in the independent sample [N=34 24]), and summary-level data on AMI (5 948 AMI cases, 355 246 controls from UK Biobank) and HF (47 309 cases and 930 014 controls from the HERMES consortium).

There was no evidence of a causal effect of circulating cTnI on AMI (Inverse Variance Weighted (IVW) odds ratio (OR) = 1.00, 95% Confidence Interval (CI) [0.99, 1.00]).

Bidirectional MR however suggested evidence for a potential causal effect of cTnI on HF (IVW OR = 1.20, 95% CI: [1.00, 1.45]) in addition to the well-known effect of HF on cTnI concentration (IVW effect size = 0.14, 95% CI: [0.00, 0.28]) (Table S17). Sensitivity analyses showed similar results between all methods (weighted mode, weighted median, penalized weighted median, MR-Egger), although wider confidence intervals due to low power, and Steiger filtering did not change the results (Table S17, Figures 4-5). The MR-Egger intercept was close to zero for both outcomes, giving no strong evidence of direct effects of the index variants on AMI or HF that are not mediated by cTnI (horizontal pleiotropy). However, there was some evidence for heterogeneity in the analysis of HF (heterogeneity statistic Q = 27, p-value = 0.003), but not for the analysis of AMI (Q = 9, p-value = 0.50). The heterogeneity in the HF analysis could possibly reflect horizontal pleiotropy, which would violate the instrumental variable analysis assumptions, or indicate multiple underlying mechanisms. Because HF development is commonly preceded by increase in left ventricular mass, we further investigated the association of cTnI on HF. We found a weak association of circulating cTnI on left ventricular mass (IVW effect size = 0.07, 95% CI: -0.11, 0.27), but the effect was attenuated and changed direction in sensitivity analyses (Table S17).

**Figure 4:**
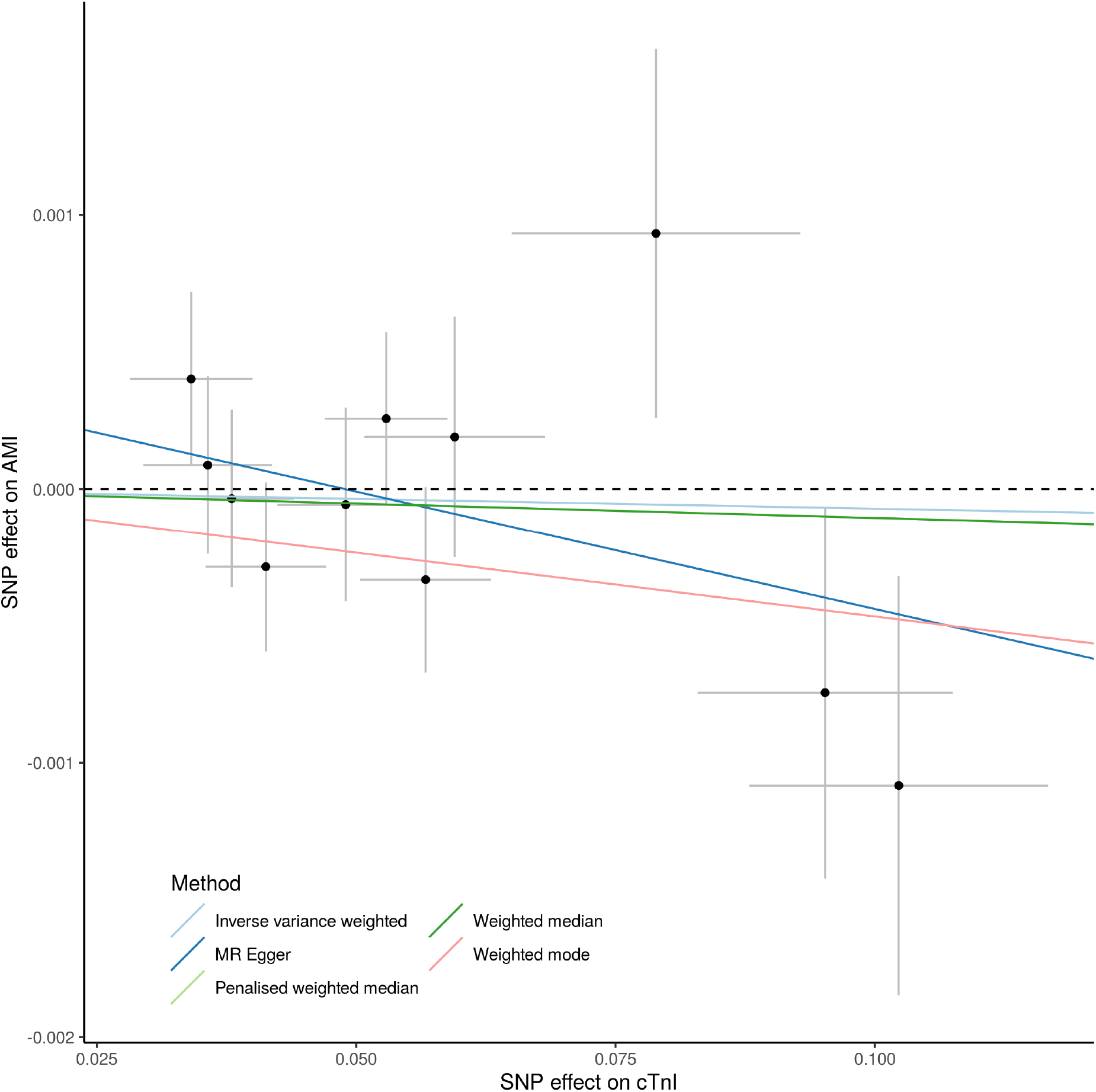
Mendelian randomization analysis of the causal effect of cTnI on AMI: The analysis was based on summary statistics from the meta-analysis of HUNT and GS:SFHS (n = 48 115) for cTnI and from a GWAS of AMI in UK Biobank from the Neale laboratory (cases/controls= 5 9948/355 246). Effect allele-cTnI associations given on x-axis and effect allele-AMI associations given on y-axis. All associations are given with 1 standard deviation error bars.

**Figure 5:**
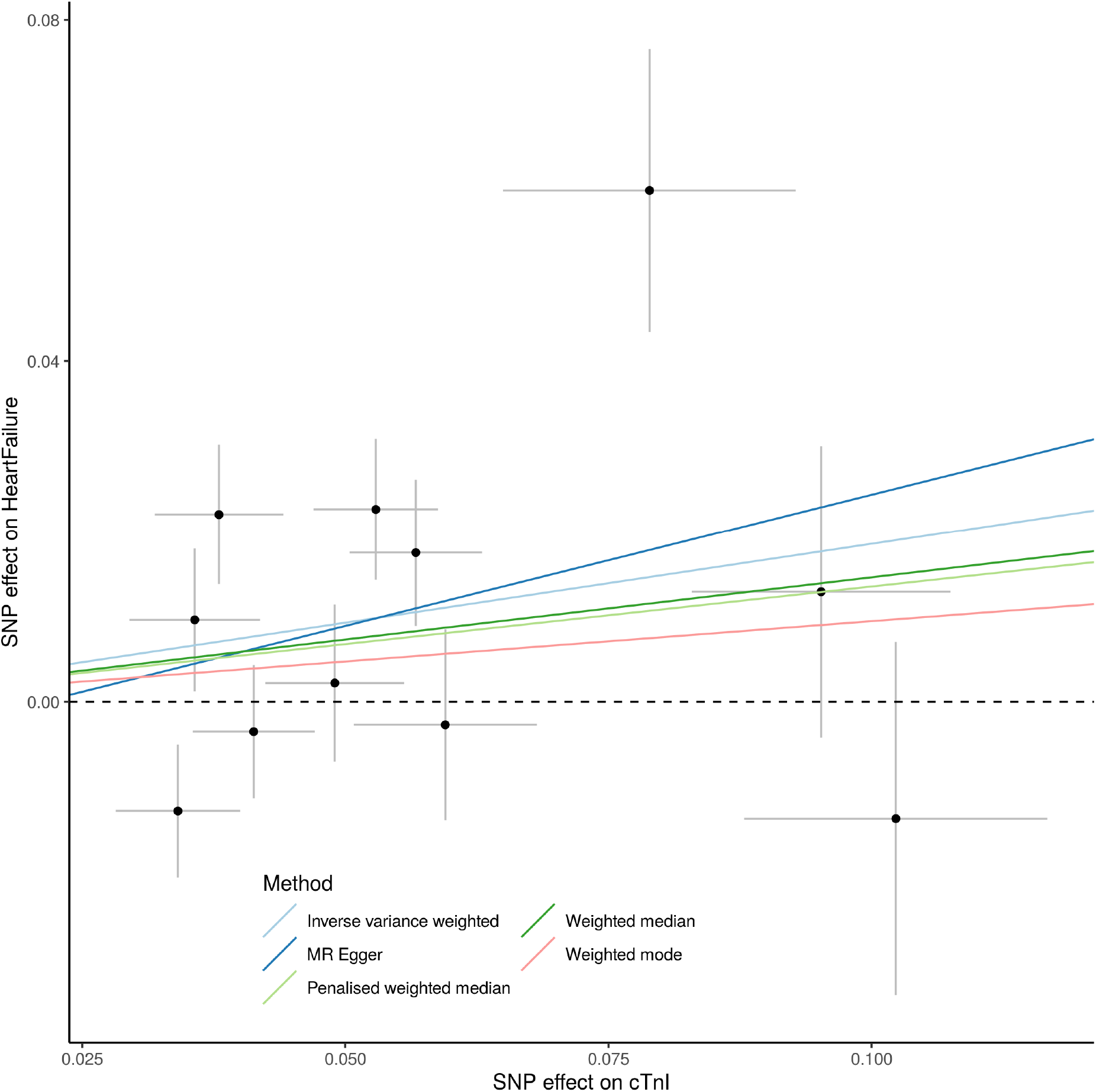
Mendelian randomization analysis of the causal effect of cTnI on HF: The analysis was based on summary statistics from the meta-analysis of HUNT and GS:SFHS (n = 48 115) for cTnI and from a meta-analysis of HF in the HERMES consortium (cases/controls = 47 309/930 014). Effect allele-cTnI associations given on x-axis and effect allele-HF associations given on y-axis. All associations are given with 1 standard deviation error bars. The outlier variant with a large effect size for both traits is rs9391746 from the HLA region.

## Discussion

Our GWAS meta-analysis of 48 115 individuals in HUNT and GS:SFHS revealed a genetic contribution to circulating protein concentrations of cTnI with associations to several biological processes. We identified 12 loci consistently associated with cTnI concentrations across both studies, confirming four of the five previously unreplicated loci(13). Furthermore, we suggest several novel loci for change in cTnI with time, confirm cTnI as a non-causal biomarker for AMI and observe indications that cTnI may have a causal effect on HF development.

Cardiac troponin I and T are important biomarkers for cardiovascular disease, and especially used to diagnose AMI in patients with symptoms of acute myocardial ischemia(1,40). Moreover, cTnI and cTnT have been found to provide strong and independent information regarding future risk for cardiac morbidity and mortality among subjects of the general population(41). Of note, while cardiac troponins are excellent markers of AMI in the acute setting, high-sensitivity troponins seem more related to incident heart failure events than to future AMI in the general population(42,43). Hence, data from several independent cohorts support a model of cardiac troponins as either directly or indirectly associated with heart failure development. This model is supported by high-sensitivity troponin concentrations being correlated with indices of structural heart disease, and most prominently left ventricular mass(42,44–46).

Using genetic data from two large cohorts from the general population (HUNT and GS:SFHS), we now provide new and additional information related to a role for cardiac troponins in cardiovascular disease. Four protein-altering index variants have previously been associated with cardiovascular health or with muscle development and injury: Firstly, a protein-altering variant in the fibrinolysis related gene *HABP2* (rs7080536) that reduces the activity of the translated protein(47), coagulation factor VII activating protease, and has been associated with carotid stenosis(48) and venous thromboembolism(49). Secondly, a protein-altering variant in *APOH* (rs1801690) that has been associated with the concentrations of creatine kinase(50), which is an unspecific biomarker for acute muscle injury, and with activated partial thromboplastin time(50), a measure of coagulation. Third, a previously reported protein-altering variant in *ANO5* (rs7481951) that has been suggested to be causal for adult-onset muscular dystrophy(51). Finally, we identified a rare protein-altering variant (rs151313792) in *FHOD3*, a gene that regulates actin assembly in cardiac sarcomeres(52) and has been associated with dilated cardiomyopathy(53) and left ventricular ejection fraction(54).

The remaining index variants were in close proximity to genes previously associated with the cardiovascular system, either through disease associations or links to cardiovascular development, structure and function: *ALPK3, FHOD3, VCL* and *CAND2* have been implicated in heart development and/or function(50,55–57), and *TNFAIP2* is central in blood vessel formation(58). Both *ALPK3* and *VCL* are associated with cardiomyopathy(4,53,55), and *CAND2* is associated with atrial fibrillation(59) and also creatine kinase concentrations(50). Furthermore, the *KLKB1*/*CYP4V2* locus has been associated with atherosclerosis(60), and the *KLKB1* and *F12* genes are both involved in several proteolytic reaction cascades in the cardiovascular system(61). These cascades are associated with coagulation/fibrinolysis, blood pressure regulation and inflammation(61). *LMAN1* has also been associated with coagulation(62), and both *TNFAIP2* and *HLA*-*DQB1* are associated with vascular inflammation(58,63). Both coagulation and atherosclerosis are pathobiological components crucial to the progress of acute exacerbations of coronary artery disease, and blood pressure regulation will impact left ventricular mass, and this is therefore comparable with previous data on cardiac troponins and left ventricular structure and function. The *F12*/*GRK6* locus has additionally been associated with chronic kidney disease(64). Several of the protein-altering and regulatory region variants that we identified had previously also been linked to some of the above-mentioned processes(47–51,60,65,66). Excluding individuals with impaired kidney function or a history of diabetes, atrial fibrillation and flutter, cardiomyopathy or HF, did not attenuate the effect sizes of any of the 12 index variants, supporting that the genetic associations with cTnI concentrations were not driven by these clinical endpoints.

Fine-mapping of the cTnI associated loci pointed to additional protein-altering and regulatory region variants, and further indicated two separate causal signals in each of the two most significant cTnI loci. A rare *ANO5* frameshift substitution (rs137854521) was possibly driving the second independent signal in the most significant locus as indicated by the fine-mapping analysis. A variant in another credible set was located in the *F12* 5’ untranslated region (UTR) and is associated with coagulation factor XII deficiency (rs1801020, r^2^_HUNT_ = 0.99 with rs2731672). The protein-altering variant in *HABP2* was alone in its credible set, providing additional support for its contribution to circulating cTnI concentrations in the general population. On the other hand, a large credible set from the barely significant HLA region contained only intergenic variants, none of which were the index variant, weakening the evidence for a role of this specific variant in troponin biology. This is not surprising, given that variants in this region are known to be difficult to impute and interpret(67).

With a small number of significant cTnI loci, the DEPICT analysis was limited by the variant database. We filtered on variants in the DEPICT database so that all cTnI loci were represented, but with the cost that the results were not necessarily based on the most significant variants within the loci. Despite this limitation, we found that the expression of genes in cTnI loci with p-value < 10^−5^ was most significantly enriched in the heart (particularly in the ventricles), which is as expected given that cTnI is only expressed in cardiac myocytes, which is also the basis for the excellent performance of cardiac troponins as cardiovascular biomarkers(68). Additionally, DEPICT prioritized several genes in the cTnI loci that were associated with heart development, structure and function, and with cardiovascular diseases: *NRAP* is involved in sarcomere assembly during cardiomyocyte development(69) and is associated with familial cardiomyopathies(70), while *CASP7* is involved in activating apoptosis(71) and is associated with cardiac electrophysiology(71). Both are located in the *HABP2* locus. Likewise, *SYNPO2L*, which is involved in sarcomeric signal transduction(72) and cardiac development(37), is located in the second most significant locus on chromosome 10, but further away from the index variant than *VCL*. The associations with sarcomere structure and function are plausible based on the biology as cardiac troponins are essential components of the sarcomere.

While we identified protein-altering variants in more than half of the loci, several statistically significant variants were located outside of the coding regions. Non-coding variants might however have regulatory functions, and we therefore investigated their associations with variation in gene expression across 49 tissues. Four cTnI loci with non-coding index variants were highly colocalized with the expression signals for a nearby gene in at least one tissue, including the expression of coagulation factors in arteries and liver, and a protein degradation activator in blood. The colocalization with expression signals for adenosine kinase was also quite strong in the aorta. This protein has been implicated in cardiomyocyte microtubule dynamics and cardiac adaption to high blood pressure and is found to attenuate cardiac hypertrophy(73). These findings corroborate the role of these loci in cTnI biology, possibly in relation to the response to cardiac injury and myocardial stress, and to the breakdown of circulating cTnI.

The single-variant phenome-wide association tests in the UK Biobank indicated some level of pleiotropy of cTnI variants: Seven of 12 index variants were associated with other traits than cTnI. These traits included cardiovascular diseases, waist-hip ratio, standing height, aspartate aminotransferase 1 concentrations and basal metabolic rate. Waist-hip ratio has been linked to future risk of cardiovascular outcomes(74), standing height correlates with blood pressure, cardiac output and vascular resistance(75), and the basal metabolic rate could potentially affect cTnI degradation. Aspartate aminotransferase 1 is an enzyme released from the heart in patients with AMI(76), but is also related to amino acid metabolism in the liver and is used as an unspecific biomarker for liver cell damage(77). This enzyme could thus both be related to cardiovascular disease and possibly to degradation and clearance of circulating cTnI in the liver. However, there were no associations between cTnI variants and the more liver-specific enzyme alanine aminotransferase 1, which could indicate that the association is related to the cardiovascular system, rather than to liver damage. Hence, the mechanism whereby aspartate aminotransferase 1 influences circulating cTnI concentrations will require additional studies. The identified HLA region variant in our study was mainly associated with autoimmune and inflammatory diseases, and this is relevant for cardiovascular diseases(78), but more studies are needed, especially since interpretation of specific HLA region variants is difficult.

The PRS combining the 12 cTnI index variants explained 1.2% of variation in cTnI concentrations in an independent sample from HUNT. In UK Biobank it was associated with some of the same traits as the single variants, which is not surprising: standing height, basal metabolic rate, aspartate aminotransferase 1 concentrations and cardiac arrhythmias, including atrial fibrillation and flutter.

Furthermore, the PRS was associated with whole body fat-free mass and whole-body water mass which was not seen for any single variant. By performing two-sample MR, we confirmed a non-causal role of cTnI in AMI. In contrast, we found indications of a potential causal effect of cTnI in HF development, although the estimate was imprecise. This was surprising given the current opinion of cTnI as a passive biomarker of cardiac injury(68), but in line with some animal studies suggesting that circulating cTnI can trigger immune responses causing HF(10,11). Follow-up analyses of left ventricular mass, which is known to increase during HF development, however, did not support this finding.

In conclusion, our study highlights diverse genetic determinants of cTnI concentration in the general population and suggests cTnI as a non-causal biomarker for AMI and HF development in the general population. Furthermore, it suggests several novel loci for change in cTnI over time, although future studies are required to validate these findings and to explore possible mechanisms.

## Data Availability

Data generated or analyzed during this study are available from the corresponding authors upon reasonable request.

## Supplemental Data

Supplemental Data include two figures and 17 tables.

## Declaration of interests

TO reports research support and honoraria from Abbott Diagnostics and Roche Diagnostics and honoraria from Siemens Healthineers, manufacturers of high sensitivity cardiac troponin assays. The other authors declare no competing interests.

## Acknowledgments

The Trøndelag Health Study (The HUNT Study) is a collaboration between HUNT Research Center (Faculty of Medicine and Health Sciences, NTNU, Norwegian University of Science and Technology), Trøndelag County Council, Central Norway Regional Health Authority, and the Norwegian Institute of Public Health. The genotyping in HUNT was financed by the National Institutes of Health; University of Michigan; the Research Council of Norway; the Liaison Committee for Education, Research and Innovation in Central Norway; and the Joint Research Committee between St Olav’s hospital and the Faculty of Medicine and Health Sciences, NTNU. The K.G. Jebsen Center for Genetic Epidemiology is funded by Stiftelsen Kristian Gerhard Jebsen; Faculty of Medicine and Health Sciences, NTNU; The Liaison Committee for education, research and innovation in Central Norway; and the Joint Research Committee between St. Olavs Hospital and the Faculty of Medicine and Health Sciences, NTNU. The cardiac troponin I analyses were sponsored by Abbott Diagnostics.

## Web Resources

Minimac4, https://genome.sph.umich.edu/wiki/Minimac4

The GTEx Portal, https://www.gtexportal.org

The Neale Laboratory,http://www.nealelab.is/ukbiobank

The Cardiovascular Disease Knowledge Portal, http://www.broadcvdi.org

## Author contributions

M.R.M analyzed the data and wrote the first draft of the manuscript. H.Røsjø contributed to the writing of the manuscript with knowledge on cardiac biology and cardiac troponin measurements. A.R, M.N.L, S.E.G, A.F.H, B.N.W, S.A.G.T, J.LF, H.Rasheed, L.F.T, W.Z, N.A., L.G.F, J.B.N and C.H contributed to analyses. B.M.B. and T.O. conceived and designed the study. All authors interpreted results and revised the paper.

## Notes

### Author Declarations

Participation in HUNT is based on informed consent, and the study has been approved by the Norwegian Data Protection Authority and the Regional Committee for Medical and Health Research Ethics in Central Norway (REK Reference number: 2015/2294). The GS:SFHS obtained written informed consent from all participants and received ethical approval from the National Health Service Tayside Committee on Medical Research Ethics (REC Reference number: 05/S1401/89). This study was covered by the ethics approval for UK Biobank studies (application 24460) from the NHS National Research Ethics Service on 17th June 2011 (Ref 11/NW/0382) and extended on 10th May 2016 (Ref 16/NW/0274).

### Summary of Updates

Data on left ventricular mass added to sensitivity analyses; results and discussion revised according to additional analyses; Additional authors added

