## Supplemental Figures S1, S2. Supplemental Tables S2-S4, S7, S8, S11, S13-S17 for "Using human genetics to understand the causes and consequences of circulating cardiac troponin I in the general population"

### Supplemental Data

Figures S1, S2
Tables S2-S4, S7, S8, S11, S13-S17

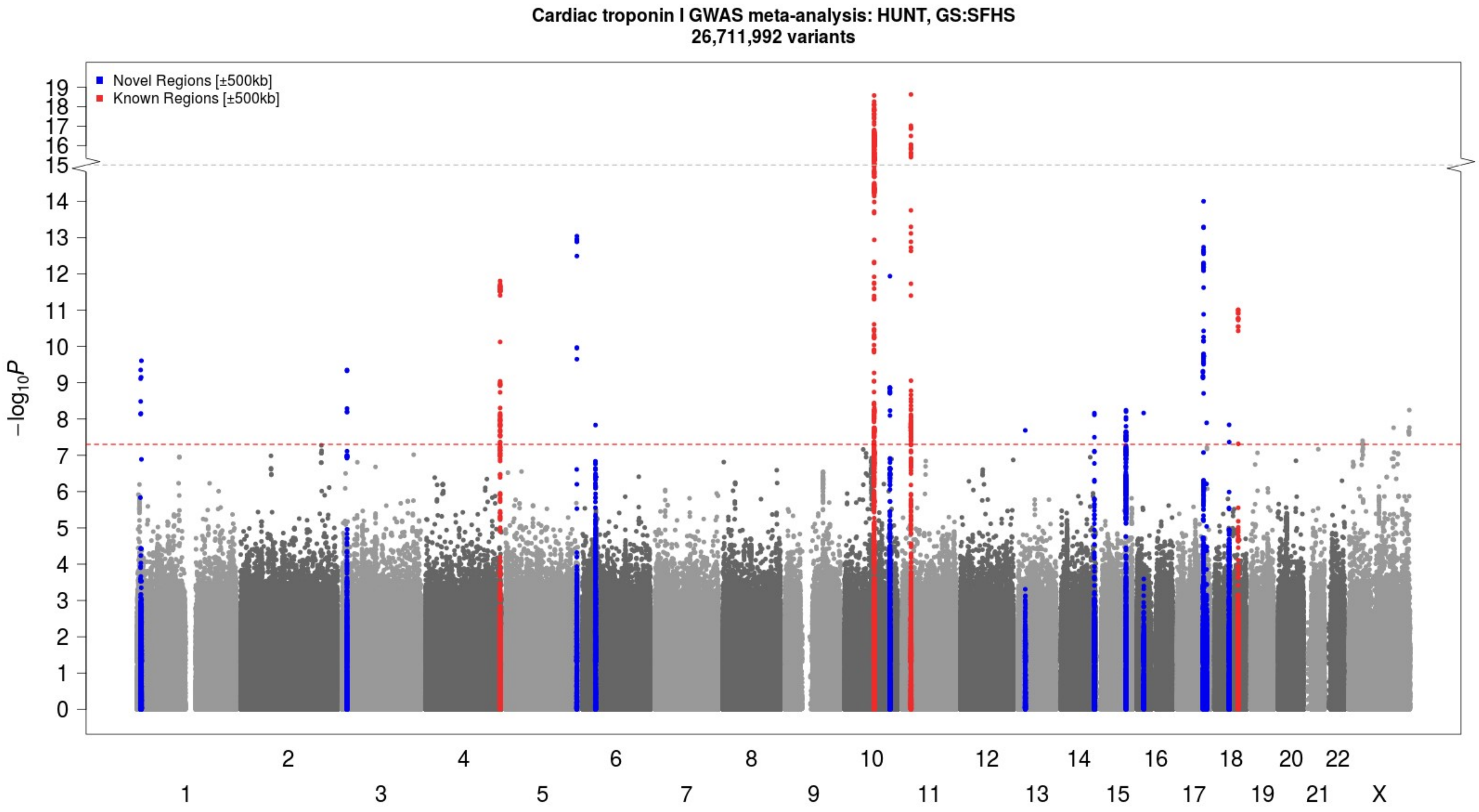

***Figure S1:*** *Manhattan plot: Genomic loci associated (p-value < 5×10$^{-8}$ ) with cTnI concentration in HUNT + GS:SFHS GWAS meta-analysis. Chromosomes are given on the x-axis and -log(p-value) for the genetic variants are given on the y-axis.*

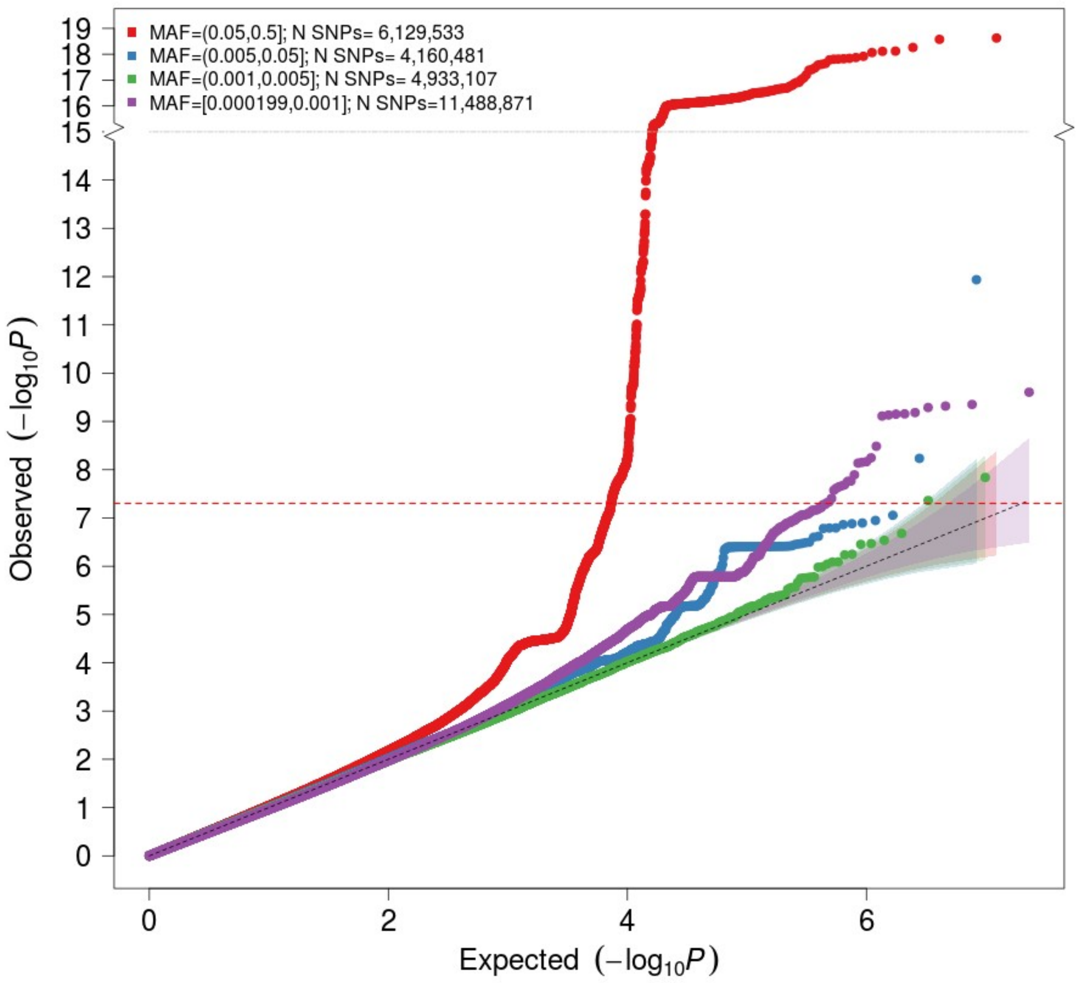

MAF=(0.05,0.5]; N SNPs= 6,129,533
MAF=(0.005,0.05]; N SNPs= 4,160,481
MAF=(0.001,0.005]; N SNPs= 4,933,107
MAF=[0.000199,0.001]; N SNPs=11,488,871
Observed (−log10P)
Expected (−log10P)

*Figure S2: Quantile-quantile plot for HUNT + GS:SFHS GWAS meta-analysis summary statistics. The inflation factor | = 1.02 (based on single nucleotide variants with MAF > 0.01).*

***Table S2:** Study Descriptives: cTnI concentration*

| | | | cTnI concentration (ng/L) | | | | Age (years) | | | Sex (% male/female) |
|---|---|---|---|---|---|---|---|---|---|---|
| | **Min** | **Max** | **IQR** | **Median** | **N** | **Min** | **Max** | **IQR** | **Median** | |
| **HUNT** | 0 | 850.28 | 1.4, 4.4 | 2.5 | 29 839 | 20 | 103 | 47, 70 | 59 | 45/55 |
| **GS:SFHS** | 0.6 | 1004.5 | 0.6, 3.1 | 1.9 | 18 276 | 18 | 99 | 37, 59 | 49 | 42/58 |

***Table S3**: Study Descriptives: Change in cTnI over time (HUNT)*

| cTnI difference / follow-up time (ng/L*years) | | | | | Age at follow-up start (years) | | | | Follow-up time (years, months) | | | | Sex (% male/female) |
|---|---|---|---|---|---|---|---|---|---|---|---|---|---|
| **Min** | **Max** | **IQR** | **Median** | **N** | **Min** | **Max** | **IQR** | **Median** | **Min** | **Max** | **IQR** | **Median** | |
| -51.77 | 38.15 | -0.18, -0.01 | -0.10 | 5 178 | 20 | 94.5 | 36.6, 63.6 | 48.6 | 9.9 | 11.9 | 10.5, 11.1 | 11.7 | 45/55 |

***Table S4**: Sensitivity analysis in HUNT*

| | | Reference (no exclusions) | | | Excluding 5 253 individuals with comorbidities | | |
|---|---|---|---|---|---|---|---|
| **rsID** | **Chr:Pos:Ref:Alt** | **Effect Size** | **SE** | **P-value** | **Effect Size** | **SE** | **P-value** |
| rs7650482 | 3:12800305:A:G | 0.04 | 0.01 | $1\times10^{-7}$ | 0.04 | 0.01 | $1\times10^{-5}$ |
| rs12331618 | 4:186218785:A:G | 0.03 | 0.01 | $7\times10^{-5}$ | 0.03 | 0.01 | $9\times10^{-5}$ |
| rs2731672 | 5:177415473:T:C | 0.05 | 0.01 | $1\times10^{-8}$ | 0.06 | 0.02 | $2\times10^{-9}$ |

| | | | | | | | |
|---|---|---|---|---|---|---|---|
| rs9391746 | 6:32715903:G:T | 0.09 | 0.02 | $7\times10^{-6}$ | 0.08 | 0.02 | $3\times10^{-4}$ |
| rs2270552 | 10:74103992:C:T | -0.06 | 0.01 | $9\times10^{-12}$ | -0.05 | 0.01 | $8\times10^{-9}$ |
| rs7080536 | 10:113588287:G:A | -0.10 | 0.02 | $9\times10^{-7}$ | -0.10 | 0.02 | $9\times10^{-6}$ |
| rs7481951 | 11:22250324:A:T | 0.05 | 0.01 | $3\times10^{-10}$ | 0.06 | 0.01 | $4\times10^{-11}$ |
| rs4283165 | 14:103137158:G:A | 0.03 | 0.01 | $1\times10^{-4}$ | 0.04 | 0.01 | $1\times10^{-5}$ |
| rs8024538 | 15:84825188:A:T | 0.03 | 0.01 | $2\times10^{-4}$ | 0.03 | 0.01 | $1\times10^{-4}$ |
| rs1801690 | 17:66212167:C:G | 0.10 | 0.02 | $5\times10^{-11}$ | 0.12 | 0.02 | $7\times10^{-12}$ |
| rs151313792 | 18:36681510:G:A | 0.32 | 0.06 | $2\times10^{-7}$ | 0.31 | 0.07 | $6\times10^{-6}$ |
| rs12604424 | 18:59353974:T:C | 0.04 | 0.01 | $3\times10^{-3}$ | 0.05 | 0.01 | $5\times10^{-4}$ |

Genomic variants are given as rsID and chromosome (Chr), position in genome build GRCh38 (Pos), reference allele (Ref) and alternate allele (Alt). Effect sizes with standard errors (SE) correspond to the alternate allele. The comorbidities that qualified for exclusion in the sensitivity analysis were impaired kidney function or a history of heart failure, cardiomyopathy, atrial fibrillation or diabetes mellitus.

##### ***S7:*** *DEPICT gene set enriched with genes in cTnI associated genetic loci*

| Original gene set ID | Original gene set description | P-value |
|---|---|---|
| MP:0001711 | Abnormal placenta morphology | $2\times10^{-18}$ |

The p-value cut-off for cTnI associated loci was set to $1\times10^{-5}$. Results with a false discovery rate < 0.05 are excluded.

##### ***Table S8:*** *DEPICT: Tissue types with enrichment of the expression of cTnI associated genes*

| MeSH term | MeSH first level term | MeSH second level term | Nominal P-value |
|---|---|---|---|

| | | | |
|---|---|---|---|
| A07.541 | Heart | Cardiovascular system | 0.0013 |
| A07.541.560 | Heart Ventricles | Cardiovascular system | 0.0015 |
| A14.549.167 | Dentition | Stomatognathic system | 0.0023 |

The p-value cut-off for cTnI associated loci was set to to $1\times10^{-5}$.

*Table S*

***11:** Significant associations between cTnI index variants and cis-eQTLs*

| rsID | Chr:Pos:Ref:Alt | Gencode ID | Gene Symbol | P-value | Tissue |
|---|---|---|---|---|---|
| rs2270552 | 10:74103992:C:T | ENSG00000156110.13 | *ADK* | $4\times10_{20}$ | Transverse colon |
| rs7481951 | 11:22250324:A:T | ENSG00000171714.10 | *ANO5* | $1\times10_{25}$ | Transverse colon |
| rs7650482 | 3:12800305:A:G | ENSG00000213943.3 | *KRT18P17* | $9\times10_{29}$ | Testis |
| rs2270552 | 10:74103992:C:T | ENSG00000185009.12 | *AP3M1* | $2\times10_{18}$ | Sigmoid colon |
| rs2270552 | 10:74103992:C:T | ENSG00000156110.13 | *ADK* | $2\times10_{14}$ | Sigmoid colon |
| rs7481951 | 11:22250324:A:T | ENSG00000171714.10 | *ANO5* | $7\times10_{17}$ | Sigmoid colon |
| rs7481951 | 11:22250324:A:T | ENSG00000171714.10 | *ANO5* | $1\times10_{24}$ | Adipose, subcutaneous |
| rs7481951 | 11:22250324:A:T | ENSG00000171714.10 | *ANO5* | $1\times10_{20}$ | Esophagus muscularis |
| rs7481951 | 11:22250324:A:T | ENSG00000171714.10 | *ANO5* | $9\times10_{22}$ | Skin, not sun exposed suprapubic |
| rs7481951 | 11:22250324:A:T | ENSG00000171714.10 | *ANO5* | $2\times10_{23}$ | Skin, not sun exposed, lower leg |
| rs2270552 | 10:74103992:C:T | ENSG00000156110.13 | *ADK* | $2\times10_{29}$ | Lung |
| rs7481951 | 11:22250324:A:T | ENSG00000171714.10 | *ANO5* | $4\times10_{44}$ | Lung |
| rs2731672 | 5:177415473:T:C | ENSG00000131187.9 | *F12* | $2\times10_{28}$ | Liver |
| rs7481951 | 11:22250324:A:T | ENSG00000171714.10 | *ANO5* | $3\times10_{24}$ | Adipose visceral omentum |
| rs12331618 | 4:186218785:A:G | ENSG00000145476.15 | *CYP4V2* | $9\times10_{21}$ | Tibial nerve |
| rs7481951 | 11:22250324:A:T | ENSG00000171714.10 | *ANO5* | $4\times10_{77}$ | Tibial nerve |
| rs12331618 | 4:186218785:A:G | ENSG00000156110.13 | *ADK* | $6\times10_{12}$ | Tibial artery |
| rs7481951 | 11:22250324:A:T | ENSG00000171714.10 | *ANO5* | $5\times10_{68}$ | Tibial artery |
| rs7481951 | 11:22250324:A:T | ENSG00000171714.10 | *ANO5* | $1\times10_{39}$ | Aortic artery |
| rs7650482 | 3:12800305:A:G | ENSG00000144712.11 | *CAND2* | $4\times10_{43}$ | Thyroid |
| rs7481951 | 11:22250324:A:T | ENSG00000171714.10 | *ANO5* | $8\times10_{47}$ | Thyroid |

| | | | | | |
|---|---|---|---|---|---|
| rs7650482 | 3:12800305:A:G | ENSG00000213943.3 | *KRT18P17* | $1\times10^{-29}$ | Skeletal muscle |
| rs7650482 | 3:12800305:A:G | ENSG00000144712.11 | *CAND2* | $8\times10^{-69}$ | Skeletal muscle |
| rs7650482 | 3:12800305:A:G | ENSG00000272263.1 | *Antisense to CAND2* | $2\times10^{-22}$ | Skeletal muscle |

Genomic variants are given as rsID and chromosome (Chr), position in genome build GRCh38 (Pos), reference allele (Ref) and alternate allele (Alt). The variants are genome-wide significant index variants in the cTnI GWAS meta-analysis and the cis-eQTL data is from samples of European ancestry in the GTEx v8 database. All associations with a Bonferroni corrected p-value < $1\times10^{-9}$ are included in the table.

***Table S13: :** cTnI index variants that are significantly associated (p-value < $2.8\times10^{-5}$) with human diseases in the UK Biobank*

| rsID | Chr:Pos:Ref:Alt | AF | N | Trait | P-value |
|---|---|---|---|---|---|
| rs7650482 | 3:12800305:A:G | 0.65 | 14820 | Atrial fibrillation and flutter | $3\times10^{-10}$ |
| rs7650482 | 3:12800305:A:G | 0.65 | 24681 | Cardiac arrhythmias | $1\times10^{-9}$ |
| rs12331618 | 4:186218785:A:G | 0.51 | 3900 | Phlebitis and thrombophlebitis | $2\times10^{-7}$ |
| rs12331618 | 4:186218785:A:G | 0.51 | 4257 | Pulmonary heart disease | $4\times10^{-7}$ |
| rs12331618 | 4:186218785:A:G | 0.51 | 3587 | Phlebitis and thrombophlebitis of lower extremities | $8\times10^{-7}$ |
| rs9391746* | 6:32715903:G:T | 0.06 | 2293 | Psoriasis and related disorders* | $3\times10^{-33}$ |
| rs9391746* | 6:32715903:G:T | 0.06 | 2237 | Psoriasis* | $3\times10^{-33}$ |
| rs9391746* | 6:32715903:G:T | 0.06 | 1684 | Psoriasis vulgaris* | $6\times10^{-26}$ |
| rs9391746* | 6:32715903:G:T | 0.06 | 2103 | Intestinal malapsorption (non-celiac)* | $2\times10^{-25}$ |
| rs9391746* | 6:32715903:G:T | 0.06 | 1855 | Celiac disease* | $1\times10^{-24}$ |
| rs9391746* | 6:32715903:G:T | 0.06 | 708 | Psoriatic arthropathy* | $8\times10^{-10}$ |

Genomic variants are given as rsID and chromosome (Chr), position in genome build GRCh38 (Pos), reference allele (Ref) and alternate allele (Alt). Allele frequency (AF) is given for the alternate allele. The number of individuals in each analysis (N) is given per phecode.
*Findings from the HLA region

*Table S*

***14:*** *cTnI index variants that are significantly associated (p-value < $2.8\times10^{-5}$) with continuous traits and biomarkers in the UK Biobank*

| rsID | Chr:Pos:Ref:Alt | AF | N | Trait | P-value |
|---|---|---|---|---|---|
| rs7650482 | 3:12800305:A:G | 0.64 | 407 399 | Waist-hip ratio | $1\times10^{-7}$ |
| rs7650482 | 3:12800305:A:G | 0.64 | 407 289 | Standing height | $5\times10^{-7}$ |
| rs7650482 | 3:12800305:A:G | 0.64 | 407 289 | Aspartate aminotransferase 1 | $2\times10^{-8}$ |
| rs7650482 | 3:12800305:A:G | 0.64 | 407 399 | Basal metabolic rate | $6\times10^{-5}$ |
| rs27316772 | 5:177415473:T:C | 0.74 | 407 289 | Standing height | $2\times10^{-9}$ |
| rs27316772 | 5:177415473:T:C | 0.74 | 407 399 | Basal metabolic rate | $4\times10^{-49}$ |
| rs7481951 | 11:22250324:A:T | 0.59 | 407 289 | Aspartate aminotransferase 1 | $7\times10^{-42}$ |
| rs8024538 | 15:84825188:A:T | 0.53 | 407 289 | Aspartate aminotransferase 1 | $6\times10^{-12}$ |
| rs1801690 | 17:66212167:C:G | 0.06 | 407 289 | Aspartate aminotransferase 1 | $2\times10^{-19}$ |

Genomic variants are given as rsID and chromosome (Chr), position in genome build GRCh38 (Pos), reference allele (Ref) and alternate allele (Alt). Allele frequency (AF) is given for the alternate allele. The number of individuals in each analysis (N) is given per trait.

***Table S***

***15:*** *Human diseases that are significantly (p-value < $2.8\times10^{-5}$) with cTnI PRS in the UK Biobank*

| Phecode | Description | P-value | $N_{cases}$ | $N_{controls}$ |
|---|---|---|---|---|
| 427.2 | Atrial fibrillation and flutter | $5\times10^{-6}$ | 14 792 | 380 170 |
| 427.0 | Cardiac arrhythmias | $1\times10^{-5}$ | 24 629 | 380 170 |

***Table S16:*** *Continuous traits and biomarkers significantly (p-value < $2.8\times10^{-5}$) associated with cTnI PRS in the UK Biobank*

| Trait | P-value | N |
|---|---|---|
| Standing height | $6\times10_{-12}$ | 407 289 |
| Aspartate aminostransferase 1 | $2\times10^{-6}$ | 387 672 |
| Whole body fat free mass | $2\times10^{-6}$ | 401 015 |
| Whole body water mass | $5\times10^{-6}$ | 401 050 |
| Basal metabolic rate | $7\times10^{-6}$ | 401 039 |

***Table S17:*** *Evaluation of the causal effects between cTnI concentrations, HF, AMI and left ventricular (LV) mass using two-sample Mendelian Randomization*

| Exposure | Outcome | Method | $N_{variants}$ | Effect Size | SE | P-value | 95% $CI_{lower}$ | 95% $CI_{upper}$ | OR | OR 95%$CI_{lower}$ | OR 95%$CI_{upper}$ |
|---|---|---|---|---|---|---|---|---|---|---|---|
| cTnI | HF | Inverse Variance Weighted | 11 | 0.19 | 0.09 | 0.05 | 0.00 | 0.37 | 1.20 | 1.00 | 1.45 |
| cTnI | HF | MR Egger | 11 | 0.31 | 0.32 | 0.36 | -0.32 | 0.94 | 1.36 | 0.72 | 2.56 |
| cTnI | HF | Penalized weighted median | 11 | 0.14 | 0.09 | 0.14 | -0.04 | 0.31 | 1.14 | 0.96 | 1.37 |
| cTnI | HF | Weighted median | 11 | 0.15 | 0.09 | 0.11 | -0.03 | 0.33 | 1.16 | 0.97 | 1.38 |
| cTnI | HF | Weighted mode | 11 | 0.09 | 0.16 | 0.57 | -0.22 | 0.41 | 1.10 | 0.80 | 1.51 |
| HF | cTnI | Inverse Variance Weighted | 11 | 0.14 | 0.07 | 0.06 | -0.00 | 0.28 | - | - | - |
| HF | cTnI | MR Egger | 11 | 0.18 | 0.22 | 0.44 | -0.25 | 0.60 | - | - | - |
| HF | cTnI | Penalized weighted median | 11 | 0.16 | 0.06 | 0.01 | 0.04 | 0.28 | - | - | - |
| HF | cTnI | Weighted median | 11 | 0.14 | 0.06 | 0.02 | 0.01 | 0.26 | - | - | - |
| HF | cTnI | Weighted mode | 11 | 0.25 | 0.14 | 0.11 | -0.03 | 0.53 | - | - | - |
| cTnI | AMI | Inverse Variance Weighted | 11 | 0.00 | 0.00 | 0.73 | -0.01 | 0.00 | 1.00 | 0.99 | 1.00 |
| cTnI | AMI | MR Egger | 11 | -0.01 | 0.01 | 0.23 | -0.02 | 0.01 | 0.99 | 0.98 | 1.01 |

| | | | | | | | | | | | |
|---|---|---|---|---|---|---|---|---|---|---|---|
| cTnI | AMI | Penalized weighted median | 11 | 0.00 | 0.00 | 0.71 | -0.01 | 0.00 | 1.00 | 0.99 | 1.00 |
| cTnI | AMI | Weighted median | 11 | 0.00 | 0.00 | 0.71 | -0.01 | 0.00 | 1.00 | 0.99 | 1.00 |
| cTnI | AMI | Weighted mode | 11 | 0.00 | 0.01 | 0.40 | -0.01 | 0.01 | 1.00 | 0.99 | 1.01 |
| cTnI | LV mass | Inverse Variance Weighted | 11 | 0.07 | 0.10 | 0.43 | -0.11 | 0.27 | - | - | - |
| cTnI | LV mass | MR Egger | 11 | -0.12 | 0.34 | 0.72 | -0.78 | 0.54 | - | - | - |
| cTnI | LV mass | Penalized weighted median | 11 | -0.00 | 0.10 | 0.98 | -0.21 | 0.20 | - | - | - |
| cTnI | LV mass | Weighted median | 11 | 0.01 | 0.11 | 0.92 | -0.20 | 0.22 | - | - | - |
| cTnI | LV mass | Weighted mode | 11 | -0.07 | 0.15 | 0.66 | -0.36 | 0.22 | - | - | - |

Effect size (approximating 1 standard deviation of the distribution of the rank transformed cTnI concentrations) with standard error (SE) and 95% confidence interval (CI) for the causal effect of the exposures on the outcomes is given per method. Odds ratios (OR) with 95% CI is given for the binary outcomes.
